# Pharmacogenomics of Coronary Artery Response to Intravenous Gamma Globulin in Kawasaki Disease

**DOI:** 10.1101/2024.01.30.24301800

**Authors:** Sadeep Shrestha, Howard W Wiener, Sabrina Chowdhury, Hidemi Kajimoto, Vinodh Srinivasasainagendra, Olga A Mamaeva, Ujval N Brahmbhatt, Dolena Ledee, Yung Lau, Luz A Padilla, Jake Chen, Nagib Dahdah, Hemant K Tiwari, Michael A Portman

## Abstract

**Background:** Kawasaki disease (KD) is a multisystem inflammatory illness of infants and young children that can result in acute vasculitis. The pathological walls of afflicted coronary arteries show propensity for forming thrombosis and aneurysms. The mechanism of coronary artery aneurysms (CAA) despite intravenous gamma globulin (IVIG) treatment is not known.

**Methods:** We performed a Whole Genome Sequencing (WGS) association analysis in a racially diverse cohort of KD patients treated with IVIG, both using AHA guidelines. We defined coronary aneurysm (CAA) (N = 234) as coronary z>2.5 and large coronary aneurysm (CAA/L) (N = 92) as z>5.0. We conducted logistic regression models to examine the association of genetic variants with CAA/L during acute KD and with persistence >6 weeks using an additive model between cases and 238 controls with no CAA. We adjusted for age, gender and three principal components of genetic ancestry. We performed functional mapping and annotation (FUMA) analysis and further assessed the predictive risk score of genomic risk loci using the area under the receiver operating characteristic curve (AUC).

**Results:** The top significant variants associated with CAA/L were in the intergenic regions (rs62154092 p<6.32E-08 most significant). Variants in *SMAT4, LOC100127*, *PTPRD, TCAF2* and *KLRC2* were the most significant non-intergenic SNPs. FUMA identified 12 genomic risk loci with eQTL or chromatin interactions mapped to 48 genes. Of these *NDUFA5* has been implicated in KD CAA and *MICU* and *ZMAT4* has potential functional implications. Genetic risk score using these 12 genomic risk loci yielded an AUC of 0.86.

**Conclusions:** This pharmacogenomics study provides insights into the pathogenesis of CAA/L in IVIG-treated KD patients. We have identified multiple novel SNPs associated with CAA/L and related genes with potential functional implications. The study shows that genomics can help define the cause of CAA/L to guide management and improve risk stratification of KD patients.

## Background

Kawasaki disease (KD) is a life-threatening acute vasculitis that diffusely affects multiple organ systems in children. Coronary artery dilatations and aneurysms can occur and represent the most serious KD complications ^1,2^ For most patients, KD is self-limited and lacks the chronic nature of other autoimmune diseases; however, the pathological walls of afflicted vessels show a propensity for forming thrombosis and aneurysms.^3^ If untreated or treatment fails, the vasculitis can lead to coronary aneurysm or thrombosis in 20–25% of cases, potentially resulting in ischemic heart disease, myocardial infarction, or death.^4,5^ Clinical trials conducted in the 1980s and 1990s showed that IVIG treatment dramatically reduced occurrence of persistent CAA defined, primarily by Japanese Ministry of Health criteria.^6,7^ These criteria stated that coronary artery diameters ≥3 mm in children <5 years and ≥4 mm in children ≥5 years were classified as abnormal. Echocardiographic detection and definition of significant CAA has dramatically improved over the past 2 to 3 decades. A 2007 study by the National Heart Lung and Blood Institute (NHLBI) showed that approximately 18 to 20% of patients had persistent CAA determined using coronary artery z scores.^8^ ^9^ The 2017 AHA guidelines adjusted the definition for CAA as z score ≥ 2.5. and ≥ 5 defines a medium to large CAA.^1^ Although these coronary abnormalities show higher prevalence in IVIG refractory patients, they can still occur in patients seemingly responsive and showing fever resolution. Most of these children with large aneurysms require daily lifelong anti-coagulation, often with twice daily painful low molecular weight heparin injections, as warfarin is difficult to maintain within therapeutic range in children.

A study based on a large-scale Japanese cohort reported that coronary events did not occur in patients with small CAA; however, 5% of patients with medium CAA and 35% of patients with large CAA had coronary events.^10^ North American studies support these data from Japan.^11^ The most severe form, giant coronary artery aneurysm (GCA), has been shown to be associated with complications such as luminal narrowing, thrombosis, and major cardiac events,^11,12^ and substantially alters quality of life for KD patients. Children with GCA require lifelong anti-coagulation, exercise restrictions, and often treatment for ischemic heart disease such as coronary stenting and/or bypass grafting.^1^ The majority of patients with GCA develop clinically important stenosis from intimal hypertrophy during the late convalescent phase.^13^ Japanese males with coronary artery involvement have a mortality rate 2.4 times higher than expected in general population^14^, but the overall impact on KD patients in the U.S. still requires definition. A recent systematic review showed that mid- to large-sized CAA provided the most significant risk factor for reducing survival of patients with KD.^15^ Mid- to large-sized CAA showed a slower recovery with worse prognosis than smaller CAA.^16^ Another study reported that the high persistence probability of mid- to large-sized CAA significantly increased the cardiovascular risk at 1 year after KD onset, when approximately two-thirds of the acute myocardial infarction cases occur.^17^

Predicting increased risk for persistent large (medium to giant) size coronary aneurysms despite IVIG treatment is clinically important for intensification of treatment and disease management. Algorithms combining clinical and lab data in Japanese populations predict risk with respect to persistent CAA.^18^ However, the Japanese algorithms show poor predictive value for risk in North American and European cohorts, ^8,19^ and fail even in some Asian populations.^20,21^ Thus, no universal biomarker or algorithm accurately predicts risk for persistent CAA in North America.^22^ Currently available data indicate that KD susceptibility and treatment response depend on an individual patient’s genetic background.^23–31^ Discrepancies among races or ethnicities also suggest that pathogenesis of KD might vary.^32,33^ To date, very few studies have focused on genetic risk factors for CAA development in KD patients. We performed the first Whole Genome Sequencing (WGS) association analysis in a cohort of KD patients in a racially diverse North American population exhibiting differences in artery aneurysm formation. We identified multiple loci associated with CAA formation among a pediatric KD population receiving IVIG that can inform on risk stratification, potentially serve as treatment response predictors and guidance toward new therapeutic targets.

## Methods

### Study populations and Primary Outcome

We performed whole genome sequencing in 504 KD patients who were diagnosed and treated with IVIG (2g/Kg on a single infusion) and aspirin,^34^ both using the American Heart Association (AHA) criteria.^1,35^ All patients included in the study had echocardiography data that assessed for coronary artery aneurysm (CAA) and were included in this study. Coronary artery internal diameters in the left main coronary artery (LMCA), left anterior descending artery (LAD), and right coronary artery (RCA) obtained by echocardiography were converted to Z scores.

Echocardiographic data were collected at baseline, at 2 weeks, and 5-6 weeks or after following fever onset. The > 5-6 week echocardiograms were assessed for persistence of the CAAs. We used the Boston z-score model^1^ and as defined by AHA, coronary abnormality or aneurysm was considered if z ≥ 2.5. We categorized positive or negative coronary artery aneurysm (CAA) occurring at any time point with z score ≥ 2.5 as “CAA”. Persistent CAA (P-CAA) was for any aneurysm z > 2.5 at ≥ 5 – 6 weeks as P-CAA We also categorized large as medium to giant coronary aneurysm for a z score ≥5.0 at any time as large CAA (CCA/L); and then persistent CCA/L (P-CCA/L) if at ≥ 5 – 6 weeks with z ≥ 5.0. Genomic comparisons were made for each of the 4 categories versus those without any aneurysm (z < 2.5).

### Whole Genome Sequencing and Variant Calling

With consent from the parents or legal guardian, whole blood or saliva was collected to extract genomic DNA, as previously described.^34^ PCR-free libraries were generated using the BGI DNBSEQ True PCR-Free platform (Beijing Genomics Institute; Guangdong, Shenzhen, China) and whole genome sequencing was performed on the MGISEQ-2000 instrument (Beijing Genomics Institute; Guangdong, Shenzhen, China) to generate 100 bp paired-end reads, as previously described.^34^ The average sequencing depth was 30x per individual. Broad Institute’s Genome Analysis Tool Kit (GATK) best practices workflow was used for quality control and informatics pre-processing of the data.

### Whole Genome Sequencing (WGS) Association - Single-Variant Analysis

Intensive quality control of the genetic data including minor allele frequency (MAF), call rate (CR), and p-values of Hardy-Weinberg equilibrium (HWE), were applied to filter uncertain SNPs as described previously,^34^ resulting in 46,718,826 variants (21,675,492 singletons) and 25,043,334 polymorphic SNPs were included in the analysis. Logistic regressions were conducted to examine the association of individual autosomal SNPs using an additive model in a case/control design, for the main outcome (medium/giant aneurysm) and the three secondary outcomes. Age, gender and three principal components (PCAs) of genetic ancestry were adjusted in the models. Quantile-quantile (QQ) plots and Manhattan plots were produced with the qqman package in R. The crude and adjusted odds ratios (ORs) and 95% confidence intervals (CIs) were also calculated for the top hits using unconditional univariate logistic regression analysis to evaluate the associations between genotypes and medium/giant aneurysm.

### Identification of Genes and Their Roles Using FUMA

Functional annotation was conducted in Functional Mapping and Annotation (FUMA) v1.3.0,^36^ using variants of interest from the WGS association analysis (*p* < 1.0 × 10^-5^ and all variants in *r*^2^ < 0.6 with them). Lead SNPs were defined from these independent statistically significant SNPs if pairwise SNPs had r^2^ < 0.1. The maximum distance between LD blocks to merge into a genomic locus was 250 kb. The genetic data of mixed population in 1000G phase3 were used as reference to estimate LD. Three methods were used to map SNPs to genes: a) physical distance (within a 10-kb window) from known protein-coding genes in the human reference assembly, b) expression quantitative trait loci (eQTL) variant mapping using,^37^ and c) 3D chromatin interaction mapping (Hi-C).^38^ Combined Annotation-Dependent Depletion (CADD) analysis^39^ with a minimum score of > 12.37 (considered to be suggestive deleterious) was used to filter the variants. Annotation of enhancers,^40^ tissue-specific expression of genes identified via Hi-C and eQTL mapping^37^ were queried in FUMA tool and Genotype Tissue Expression (GTEx) database (https://gtexportal.org/home/).

### Genetic Risk Score Computation

All genomic risk loci from FUMA were used to estimate gene risk score (GRS) by a simple risk alleles count method. Discriminative power attributable to the GRS was estimated and compared by plotting receiver operating characteristic (ROC) curves and calculating the area under the curve (AUC) for the case-control samples. The AUC compares the rates of true positives (sensitivity) and false positives (1 – specificity) and assesses the overall performance of genetic risk score models. Next, case and control status was randomly permuted 10,000 times and AUC was estimated with each pseudo case-control status. An empirical p-value (eP), which is the proportion of AUC based on randomization distribution of cases and controls that are more extreme than our observed AUC from the actual case (medium/giant aneurysm) and control (no aneurysm) status, was then calculated.

## Results

### Genetic Association Analysis Between Individual SNPs and the Risk of Large (medium/giant) Aneurysm

To identify SNPs associated with KD-associated large (medium/giant) coronary aneurysm (CCA/L), clinical data were linked in KD patients with whole genome sequencing data, nested in a clinical cohort as previously described.^34^ Basic demographics of the study population with CCA/L (N = 92) and no aneurysm (N = 276) are described in **Table 1**. Principal component analysis confirmed a good match between KD patients with CCA/L and those without any aneurysm. However, 3 PC were adjusted for in the analyses, as estimated in previous study.^34^ A quantile-quantile plot indicated that population stratification had negligible effects on the statistical results (λ genomic control = 0.955). There were several SNPs that exhibited suggestive statistical significance (p < 10^-5^) in the additive genetic model, as shown in the Manhattan plot (**Figure 1**). Of all the SNPs examined, rs62154092 in the intragenic region (nearest gene *ACTR3BP2*) was the most statistically significant (6.32E-08). Among the overall top 10 most significant SNPs, 5 SNPs (all intergenic C20_29360719, C20_29679725, C20_28616821, C20_29821717, C20_28616815) were in chromosome 20, although in different regions. All SNPs statistically significant at p <10^-4^ are listed in **Supplementary Table S1**. Among the non-intergenic SNPs, rs28730284 upstream of *KLRC2* gene was the most statistically significant (2.20 E-07) and among the top 15 non-intergenic, they were mostly intronic (rs9643846, rs9643847, rs57504215, rs60545202, rs59556769, rs73677451, rs12676292, rs4332118, rs6988966) located in *SMAT4* and others in *LOC100127* (non-coding RNA rs10276547, rs10280266), *PTPRD* (intronic rs600075, rs5896385) and *TCAF2* (intronic C7_142457900) genes. The most significant exonic SNPs (rs11259953 and rs11259954) were in *WHAMM* gene (**Supplementary Table S1**). Regional association plots with cluster of SNPs in LD in chromosomes 7, 8 and 9 are shown in **Figure 1 b-d**). Results of corresponding single SNP association with developing any CAA (N = 233), any persistent CAA (N = 145) for 2 years, and P-CCA/L for 2 years (N –=79) vs no aneurysm (N = 276) are shown in **Figure 2** and **Supplementary Table S1**.

**Figure 1.**
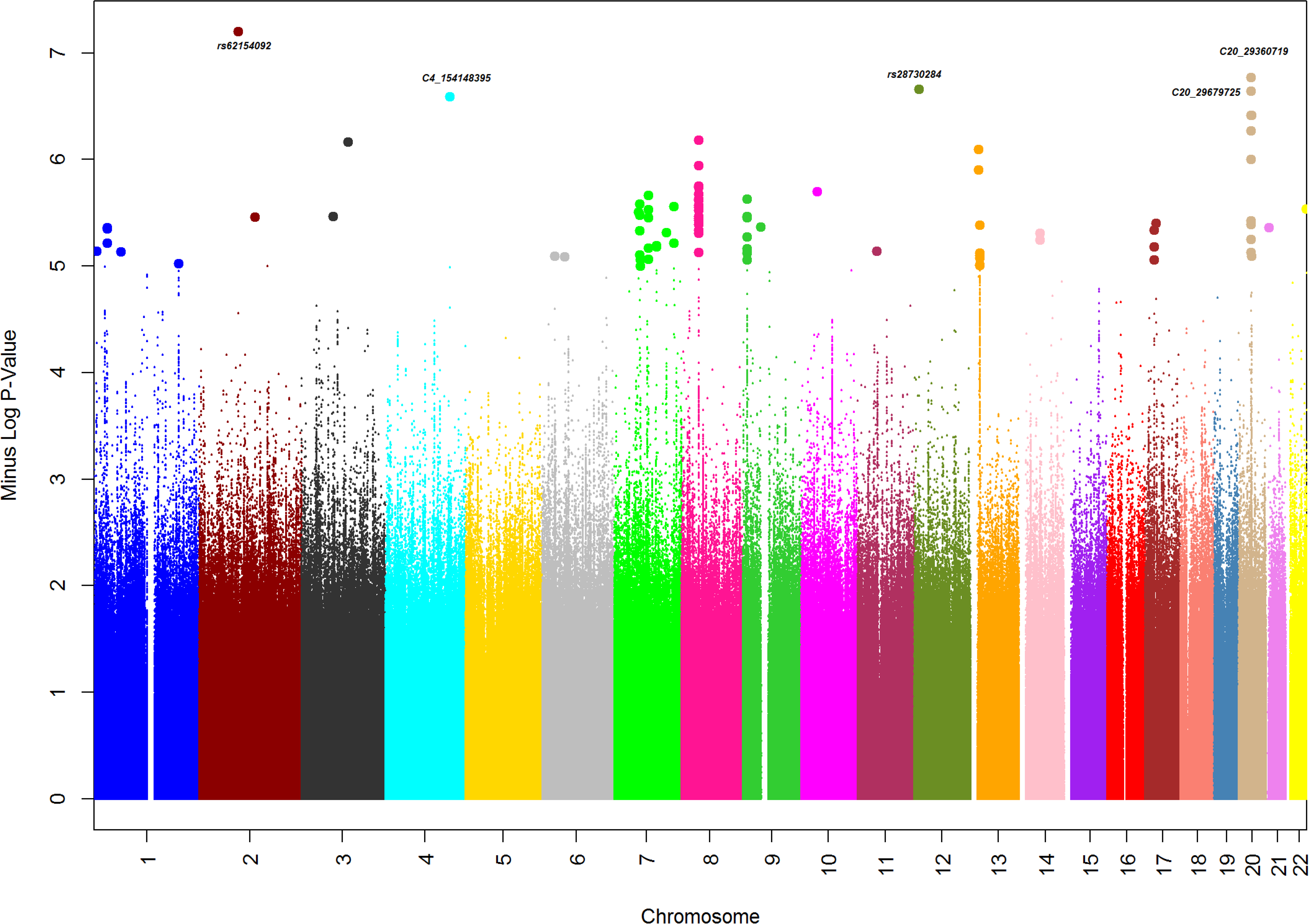
Overall and Regional Association Results. A) Manhattan Plot displaying Whole Genome Sequence Association results with - Large (medium/giant) Coronary Aneurysm (N =92) vs no Coronary Artery Aneurysm (N = 276). Negative log_10_-transformed *P* values from the logistic regression model (additive model) are plotted on the *y-*axis and the SNP genomic locations on the x-axis (colors representing different autosomal chromosomes). Locus Zoom plots for selected gene regions in B) Chromosome 7 with rs10276547 the most significant SNP in the region, C) Chromosome 8 with rs9643846 the most significant SNP in the region, D) Chromosome 9 with rs600075 the most significant SNP in the region. Vertical axis (on the left) is the –log_10_ of the p-value, the horizontal axis is the chromosomal position. Each dot represents a SNP tested for association with large coronary aneurysm. Linkage disequilibrium between the most significant SNP, listed at the top of each plot, and the other SNPs in the plot is shown by the r^2^ legend in each plot. Vertical axis (on the right) is the recombination – the site and rate are represented by red curves.

**Figure 2.**
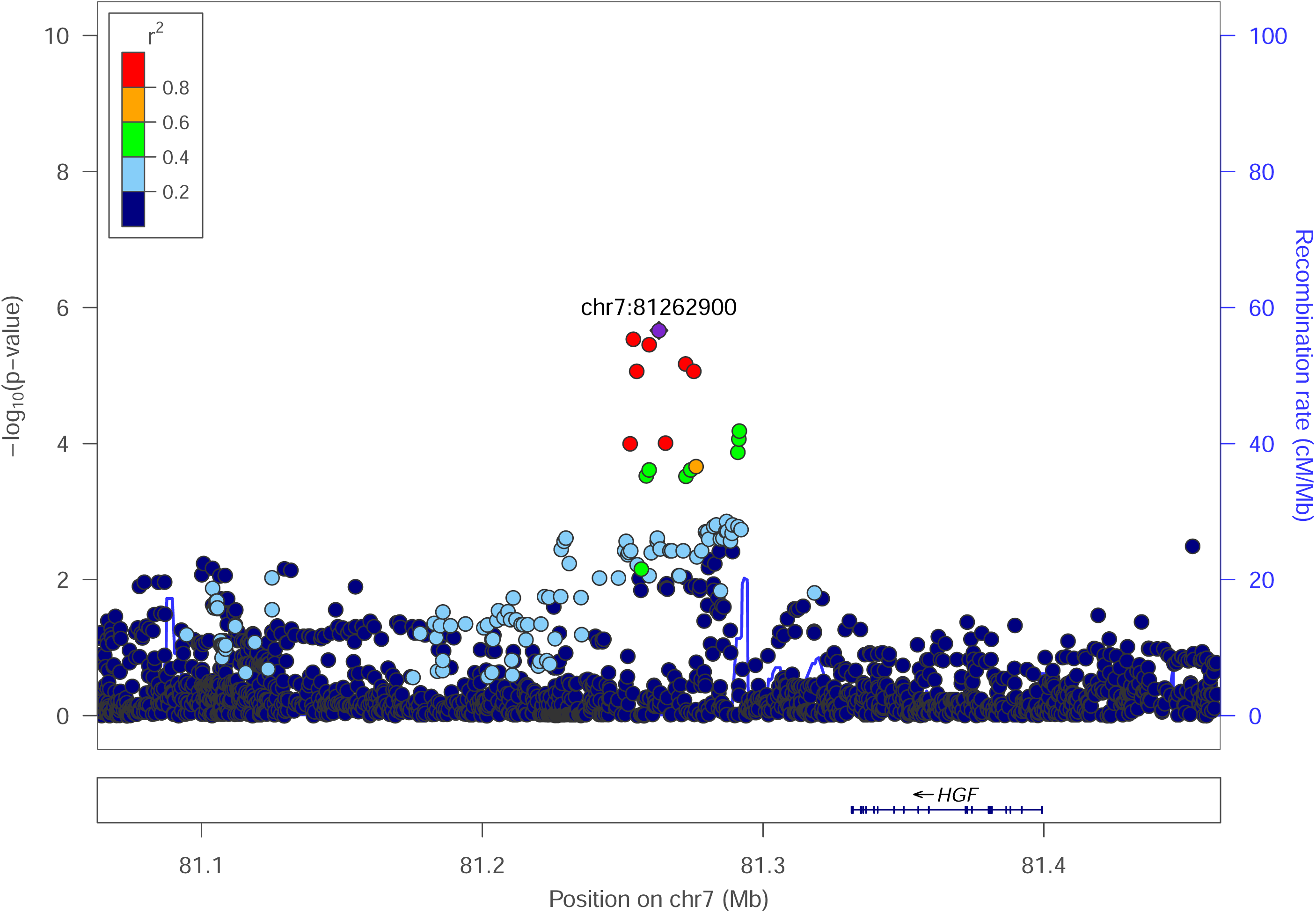
Circo plot summarizing whole genome sequence associations –. the outer plot is the association of large aneurysm (N =92), the next inner plot is the association of persistent large aneurysm (N = 79), the next inner plot is the association of any coronary aneurysm (N = 233) and the fourth inner plot is the association of persistent any coronary aneurysm vs no coronary aneurysm (for each outcome). The innermost plot indicates if SNPs were associated in 1-4 outcomes with p <10E-5.

**Table 1.**
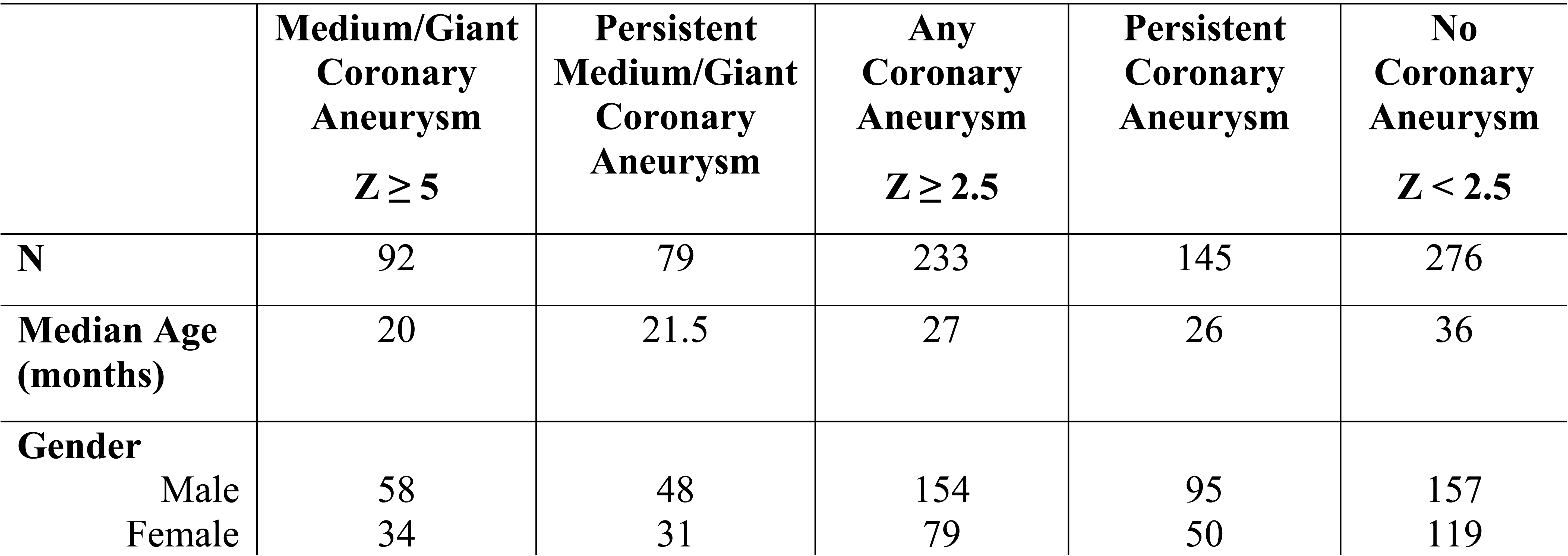
Kawasaki Disease Patients With (medium/large, any) and Without Coronary Aneurysm Included in the Whole Genome Sequencing Analyses.

### Gene mapping

Using three gene mapping strategies (position mapping, eQTL mapping and chromatin interaction mapping) in FUMA, we mapped the significant association variants (P < 10^-5^) to genes and identified 12 genomic risk loci (**Supplementary Table S2**) and 48 mapped genes associated with CCA/L (**Figure 3, Supplementary Table S3**). None of the genes were mapped by all three strategies. Three genes *NDUFA5*, *ZMAT4* and *MICU2* were mapped by physical and eQTL - *NDUFA5* is located at the chromosome 7, and its lead SNP rs34163760 is located in the intron of the gene (*P* = 4.84 × 10^−6^). An eQTL analysis showed that with the increasing number of risk alleles of rs34163760, there was a higher mRNA level of *NDUFA5* in the Esophagus. The CADD score of rs34163760 is 14.39 indicating a deleterious mutation. *ZMAT4* is located in chromosome 8, and its lead SNP rs9643846 is located in the intron of the gene (*P* = 6.57 × 10^−7^). An eQTL analysis showed that with the decreasing number of risk alleles of rs9643846, there was a higher mRNA level of *ZMAT4* in thyroid. CADD score of rs9643846 is 15 indicating a deleterious mutation. The third gene *MICU2* is located in chromosome 13, and its lead SNP rs12585631 is located in the intron of the gene (*P* = 4.11× 10^−6^). An eQTL analysis showed that with the decreasing number of risk alleles of rs12585631, there was a higher mRNA level of *MICU2* in thyroid. The CADD score of rs12585631 is 15.22 indicating a deleterious mutation. Several genes in chromosomes 4, 7 and 13 were also identified to interact with the chromatin at those sites (**Figure 3, Supplementary Table S3**).

**Figure 3.**
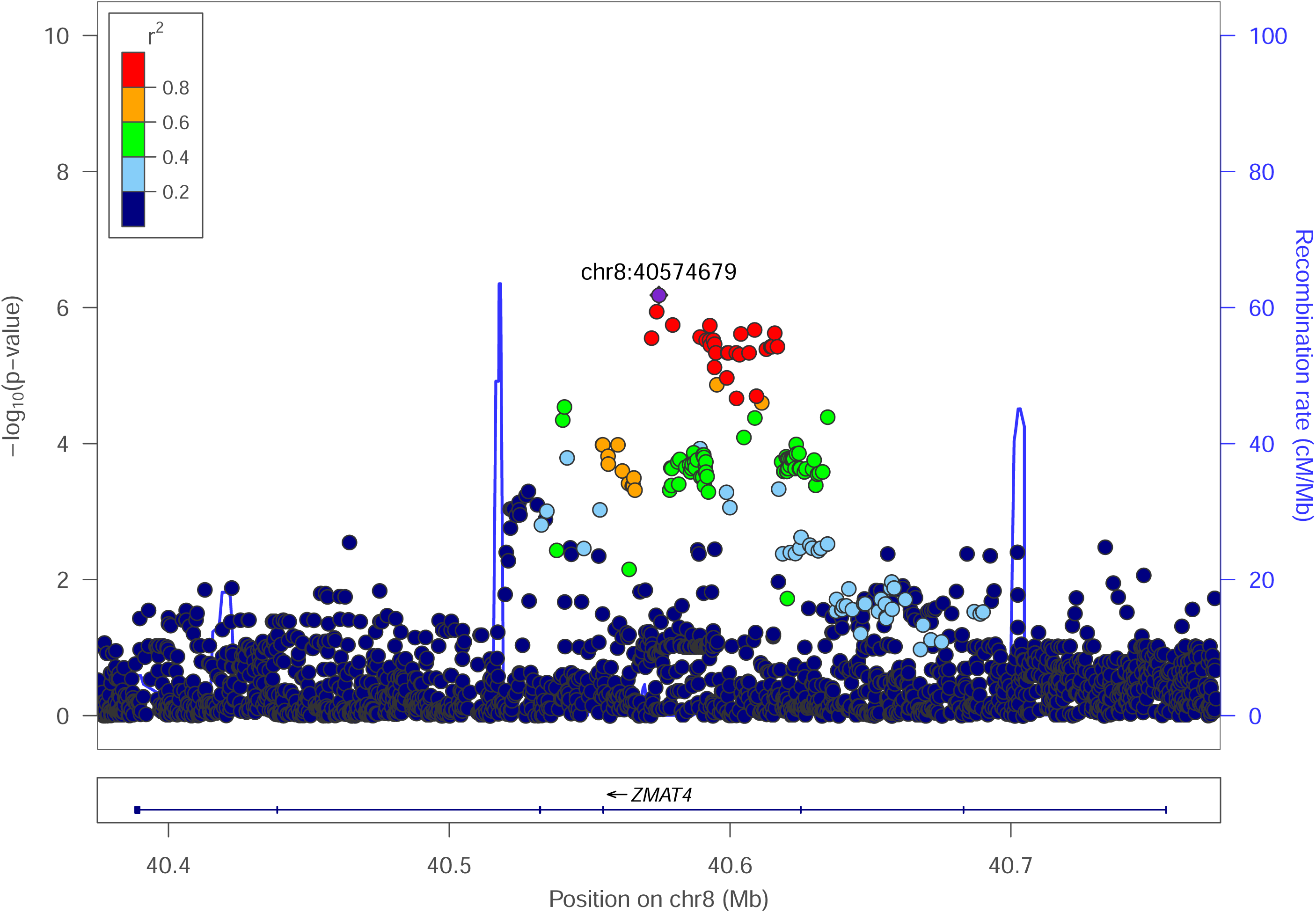
FUMA circos plots of mapped genes in genomic risk loci. - The most outer layer is the Manhattan plot (only SNPs with *P* < 0.05 are displayed). Genomic risk loci are highlighted in blue and the strength of linkage disequilibrium r^2^ between each SNP to the lead SNP is given by the following color code: red (r^2^ > 0.8), orange (r^2^ > 0.6), green (r^2^ > 0.4), blue (r^2^ > 0.2) and grey (r^2^ ≤ 0.2). Genes are mapped by 3-D chromatin interaction (orange) or eQTLs (green), or both (red). A) Circos plot for Chromosome 13 with lead SNP rs12585631, B) Circos plot for Chromosome 7 with lead SNP rs10276547, and C) Circos plot for Chromosome 4 with lead SNP rs62330192

Expression patterns of the 48 prioritized genes were estimated in 54 different tissues (**Supplementary Table S4 Supplementary Figure S1).** Several of these genes show high expression in aorta and coronary artery tissues.

### Genetic Risk Score (GRS)

Twelve genomic risk loci identified from FUMA yielded an AUC of 0.86. As shown in **Figure 4**, based on the empirical distribution of the AUC from the permutation test, eP was <0.0001, suggesting highly significant genetic risk score from the 12 genomic risk loci. For sensitivity analyses, when GRS for CCA/L was conducted separately in four specific races, AUC of 0.83, 0.78, 0.81 and 0.97 were obtained among White, Asian, Hispanic and African American KD patients.

**Figure 4.**
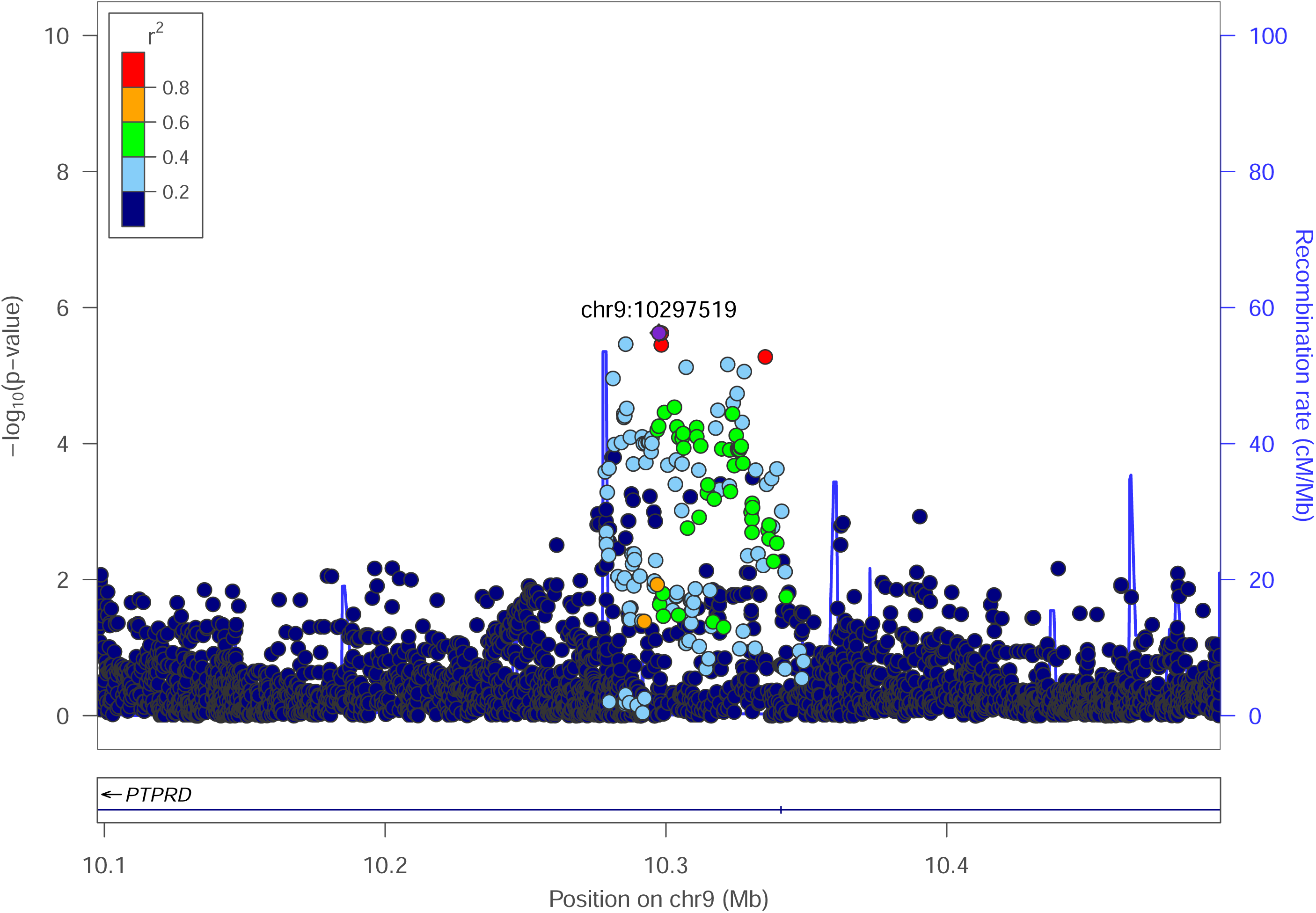
Empirical curve based on the AUC from 10,000 permutation. X-axis is the AUC value based on the 12 genomic Loci from FUMA in predicting the risk score of having a large aneurysm and Y-axis is the frequency. True AUC of 0.

## DISCUSSIONS

We analyzed a relatively large North American KD cohort using whole genome sequencing. Treatment with IVIG during the acute phase was an inclusion criterion, so lack of treatment was not a confounding factor. Additionally, persistent coronary artery aneurysm (P-CAA or P-CCA/L) should be considered a failure of IVIG therapy. As noted in Table 1, the vast majority of medium to giant coronary aneurysms persisted. We identified for the most part novel gene loci that appear to have a relationship with coronary artery aneurysm formation and persistence in KD patients. We have used this same WGS strategy to identify genes related to IVIG refractoriness as defined by AHA guidelines.^1^ Prior studies searching for CAA genetic risk variants have used either hypothesis driven strategies or genome-wide association strategy with their inherent limitations. In this study, we specifically used the newest AHA classifications, and sought to determine genetic associations with CCA/L (Z ≥ 5) as these show a lower chance for early regression than do smaller aneurysms (Z > 2.5, but < 5). However, we also found that statistical genetic associations were consistent among all coronary phenotype groups as shown in **Table 2** and **Figure 2**. This suggests that pathobiology or at least genetic risk is consistent regardless of size of the aneurysm or propensity for regression.

**Table 2.**
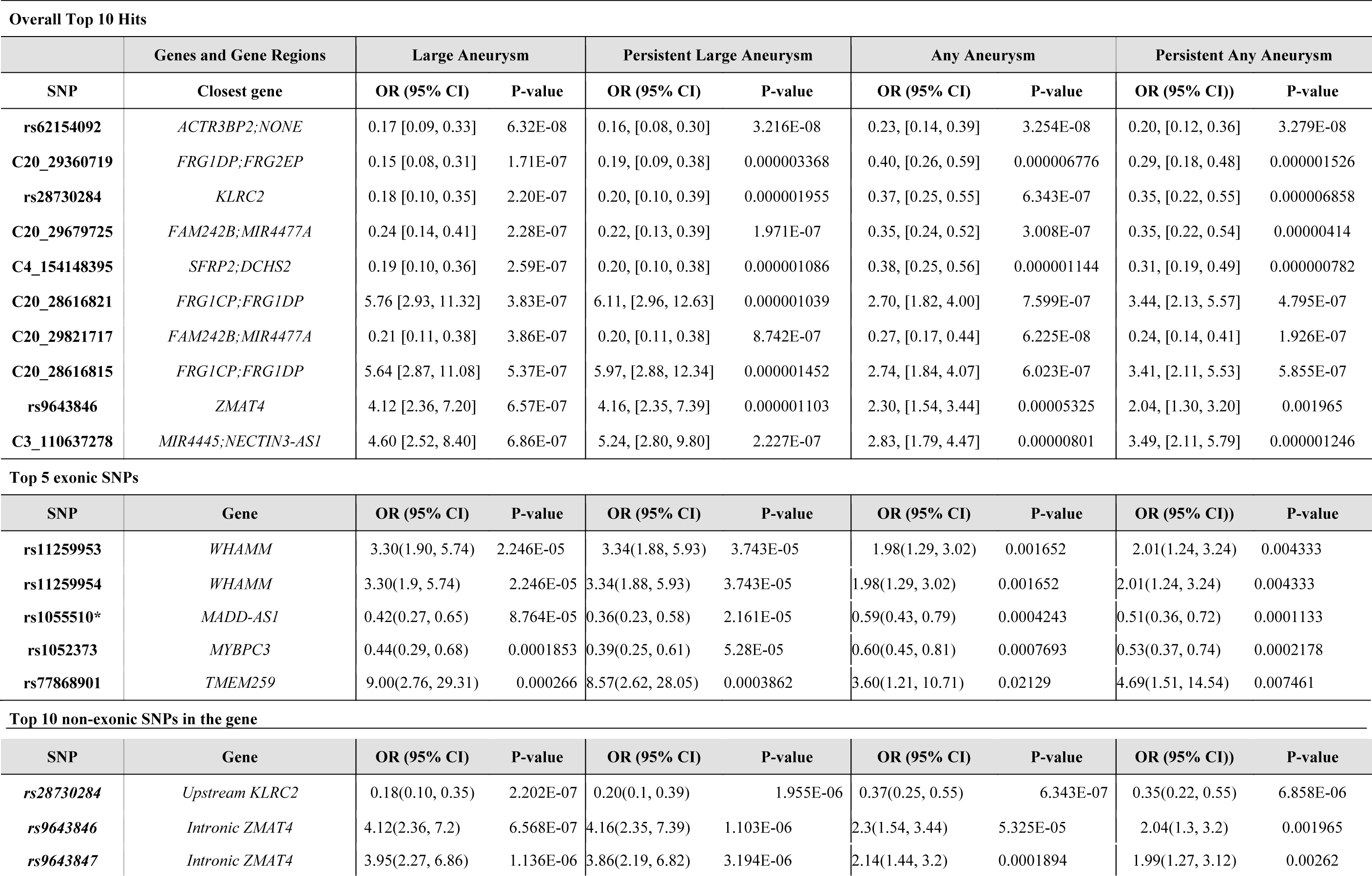
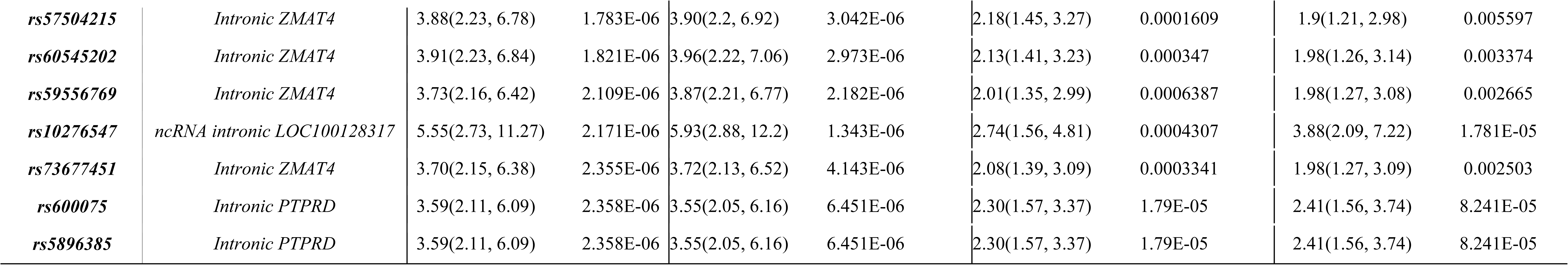
Top Genes Associated with Large (Medium/Giant) Coronary Aneurysm (CCA/L) (z ≥5), Persistent CCA/L (P-CCA/L), any coronary artery aneurysm (CAA) (z≥2.5), and persistent CAA (P-CAA)

The most significant SNP related to moderate to giant aneurysm was located in the intergenic region with the closest gene being *ACTR3BP2*, a pseudogene with unknown function. However, among the top SNPs in the gene region, rs28730284 was located just upstream of the *KLRC2* gene and has intriguing potential biological relevance to KD. The *KLRC2* gene encodes the C-type lectin NKG2C (Killer cell lectin receptor-2). Natural Killer (NK) cells mediate innate immune responses against virally infected and malignant cells.^41–43^ NK cell function such as production of proinflammatory cytokines, depends on a balance between activating and inhibiting signals triggered by multiple surface receptors, including NKG2C.^44^ Polymorphisms in *KLCR2* have been shown to influence both function and expression of NK cells. Furthermore, SNPs in *KLRC2* are associated with microvascular inflammation during renal graft transplant rejection. Importantly, all type NK cell (CD56^++^CD16^+−^, CD56^+^CD16^+^, CD56^−^CD16^+^) expression is reduced in KD patients compared to febrile or non-febrile controls, while CD56−CD16+ NK cell expression was significantly lower in IVIG-resistant patients than in the IVIG-responsive. ^45^

Multiple intronic SNPs were found in *ZMAT4* gene, which encodes the Zinc Finger Matrin-Type 4 protein. This gene was also identified by FUMA. SNPs within ZMAT4 are associated with diseases such as Spinocerebellar Ataxia and Myopia,^46^ and copy number variations are associated with hematological malignancies.^47^ Function of this particular Zinc-Finger protein remains undefined, so a potential biological role in KD would be unclear. The top two exonic SNPs were in the *WHAMM* gene. This gene encodes a protein nucleation-promoting factor that regulates the Actin-related protein 2/3 complex, but any biological relevance to KD would be highly speculative. Additionally, we found SNP (rs1052373) within the *MYBPC3* exon region as marginally significant. MYBPC3 (myosin binding protein c3) function is well established and mutations are involved in the pathology of hypertrophic cardiomyopathy.^48,49^ This particular SNP has also been associated with athletic endurance.^50^ However, any suggestion of biological relevance for these exonic SNPs to KD would be highly speculative.

Multiple SNPs were also found in regions near Facioscapulohumeral muscular dystrophy (FSHD) region-1(*FRG1DP*). FRG1 acts on upstream of FGF2, which signals activation of the AKT/ERK signaling axis in endothelial cells. Interestingly, we previously reported that this gene is associated with IVIG response in KD patients. *FRG1DP* has been linked to angiogenesis and retinal vasculpathy in FSHD patients including development of micro aneurysms. Additionally, altered expression for FRG-1 protein leads altered angiogenesis in human umbilical vein endothelial cells (HUVECs).

Using FUMA we found several genes potentially related to CAA/L in IVIG-treated KD patients. *MICU2* is a calcium sensitive regulatory subunit of the mitochondrial calcium uniporter and is important for reducing oxidative stress particularly in endothelial cells. MICU 2−/− mice exhibit abnormal cardiac diastolic relaxation but also develop abdominal aortic aneurysms, which spontaneously rupture with only modest increases in blood pressure. *NDUFA5* encodes NADH dehydrogenase [ubiquinone] 1 alpha subcomplex subunit 5, a critical component of mitochondrial respiratory complex 1, which facilitates the translocation of protons across to the mitochondrial inner membrane. A SNP in *NDUFA5* was also a top hit in a small Taiwanese KD study evaluating genetic risk for CAA formation.^51^ Thus, our findings for these two genes regulating oxidative stress response suggest that mitochondria play a role in development of CAA and should be a target for future research.^52^

We previously published a WGS pharmacogenomics analyses of IVIG response in KD using “persistent or recurrent fever” as the benchmark for IVIG resistance.^34^ Data suggests that IVIG resistance or refractoriness is a risk factor for persistent CAA. However, we did not find numbers of variants that were associated with both coronary aneurysms and IVIG resistance. This lack of genetic association uniformity between these two different outcomes suggests that their biological pathways and mechanisms may be different. However, the list of novel genes provides new insights into the pathogenesis of KD.

Genetic analyses of coronary aneurysms in KD are complicated by multiple potential confounding factors. Prior studies, which have identified numerous significant variants associated with coronary aneurysms, did not account for the IVIG therapeutic effect.^53^ ^54,55^ Clinical trials clearly show that IVIG reduces the risk of CAA.^56^ However, those previous genetic studies for the most part do not clarify whether study participants received appropriate IVIG treatment. Thus those prior cohorts could and probably do include patients whom did not receive timely IVIG. We used strict criteria for IVIG treatment in our study subjects in accordance with pharmacogenomics design. Accordingly, our results using different design and methodology did not replicate findings from prior studies such as associations with coronary aneurysms for variants in *ITPKC*^55^, *KCNN2*^54^, *NEBL* and *TUBA3C*^51^, *SLC8A1*^57^, and the matrix metalloproteinase (*MMP*) gene family^58^.

Two other studies indicated association of TIFAB^59^ and PLCB1^60^ genes. Although the same SNPs were not replicated in our study, we found several SNPs in these gene regions that were associated with CAA in our study (**Table S1**). Unlike studies predominantly using Asian populations, our cohort included 4 races (Whites, Asians, African-Americans, and Hispanics). We had fewer cases of African-Americans and Hispanics and we were underpowered to conduct race-specific analyses (**Supplementary Table S5**). However, in the main combined cohort, we adjusted for three principal components which resulted in λ genomic control = 0.955, suggesting no major confounding factors (**Supplementary Figure S2**).

We also used 12 genomic loci from FUMA to test for overall prediction of risk for developing CAA/L, a genetic predictive risk score. The AUC based on these markers is promising and the empirical risk models based on 10,000 simulated cases and controls had considerably higher AUC than theoretically achievable. Although sample size is small, all race-specific analyses also showed similar trends with high AUC. These specific markers need to be validated in different IVIG-treated KD populations; however, there is potential clinical utility in developing a point-of-care assay based on panels of genetic risk markers to predict who will develop CAA/L and/or other sequalae. Accurate prediction can assist in developing a treatment plan during the acute KD phase.^61^

The main limitation for this study is the lack of a validation cohort. This is a common limitation of clinical trials and pharmacogenomics studies of rare diseases, although our cohort is the largest reported to date with a clear IVIG treatment phenotype. Further validations as well as functional studies of these variants will be needed in the future.

In summary, using WGS we have identified several novel genes and loci, which could have a functional impact on coronary artery response to IVIG in KD. Additionally, these loci could be used in developing an important predictive risk score for persistence of coronary artery aneurysms despite IVIG treatment.

## Data Availability

The data have been deposited with links to BioProject accession number PRJNA1055092 in the NCBI BioProject database and will available per the repository guidelines (https://www.ncbi.nlm.nih.gov/bioproject/).

## Acknowledgments

The parent cohort/study and this genomic study conformed to the procedures for informed consent (parental permission) approved by institutional review boards at all sponsoring organizations and to human-experimentation guidelines set forth by the United States Department of Health and Human Services. We thank the participating patients and their parents. This study was supported by the National Institutes of Health grant **NHLBI-1R01HL146130.**

## ABREVIATIONS

AUC: Area Under the Curve
CAA: Coronary Artery Aneurysm
CAA/L: Large Coronary Artery Aneurysm
CADD: Combined Annotation-Dependent Depletion
DNA: Deoxyribonucleic Acid
eQTL: Expression Quantitative Trait Loci
FUMA: Functional Mapping and Annotation
GATK: Genome Analysis Tool Kit
GRS: Gene Risk Score
GTEx: Genotype Tissue Expression
HWE: Hardy-Weinberg equilibrium
IVIG: Intravenous Immunoglobulin
KD: Kawasaki Disease
LAD: Left Anterior Descending Artery
LD: Linkage Disequilibrium
LMCA: Left Main Coronary Artery
MAF: Minor Allele Frequency
PCA: Principal Components Analysis
QC: Quality Control
QQ: Quantile-quantile
RCA: Right Coronary Artery
ROC: Receiver Operating Characteristic
SNP: Single Nucleotide Polymorphism
WGS: Whole Genome Sequencing

## Supplementary Figures

**Supplementary Figure S1.**
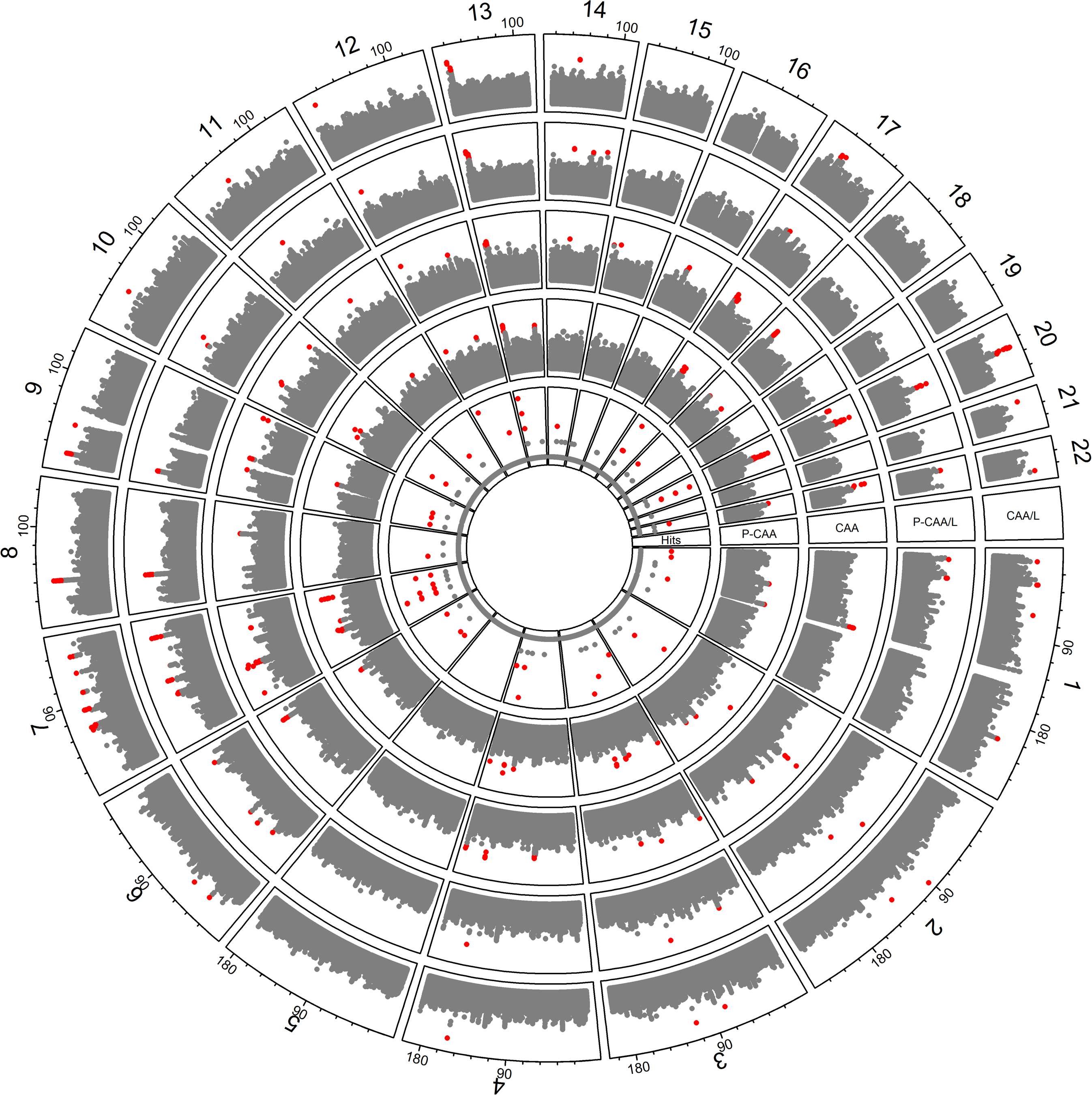
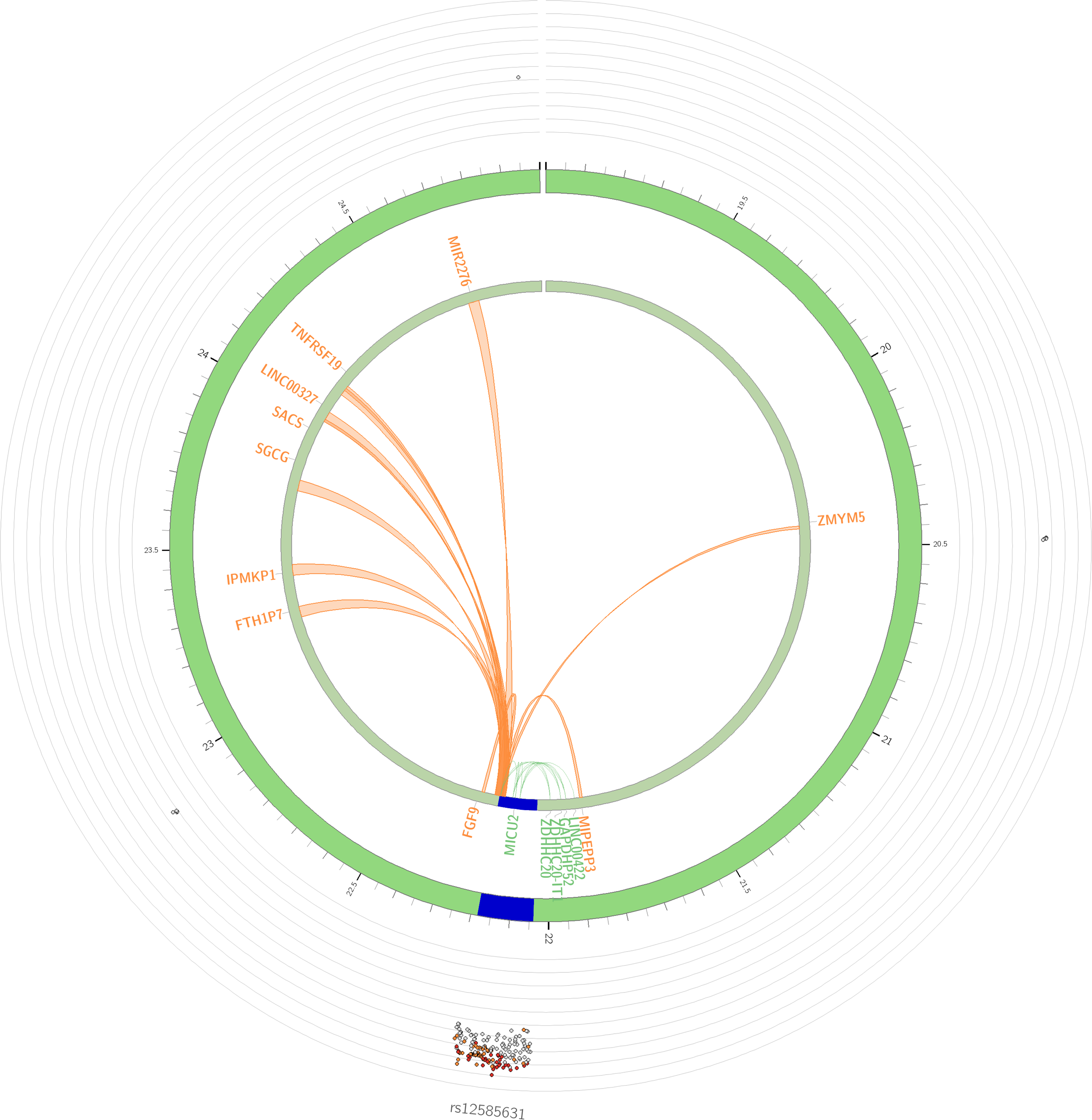
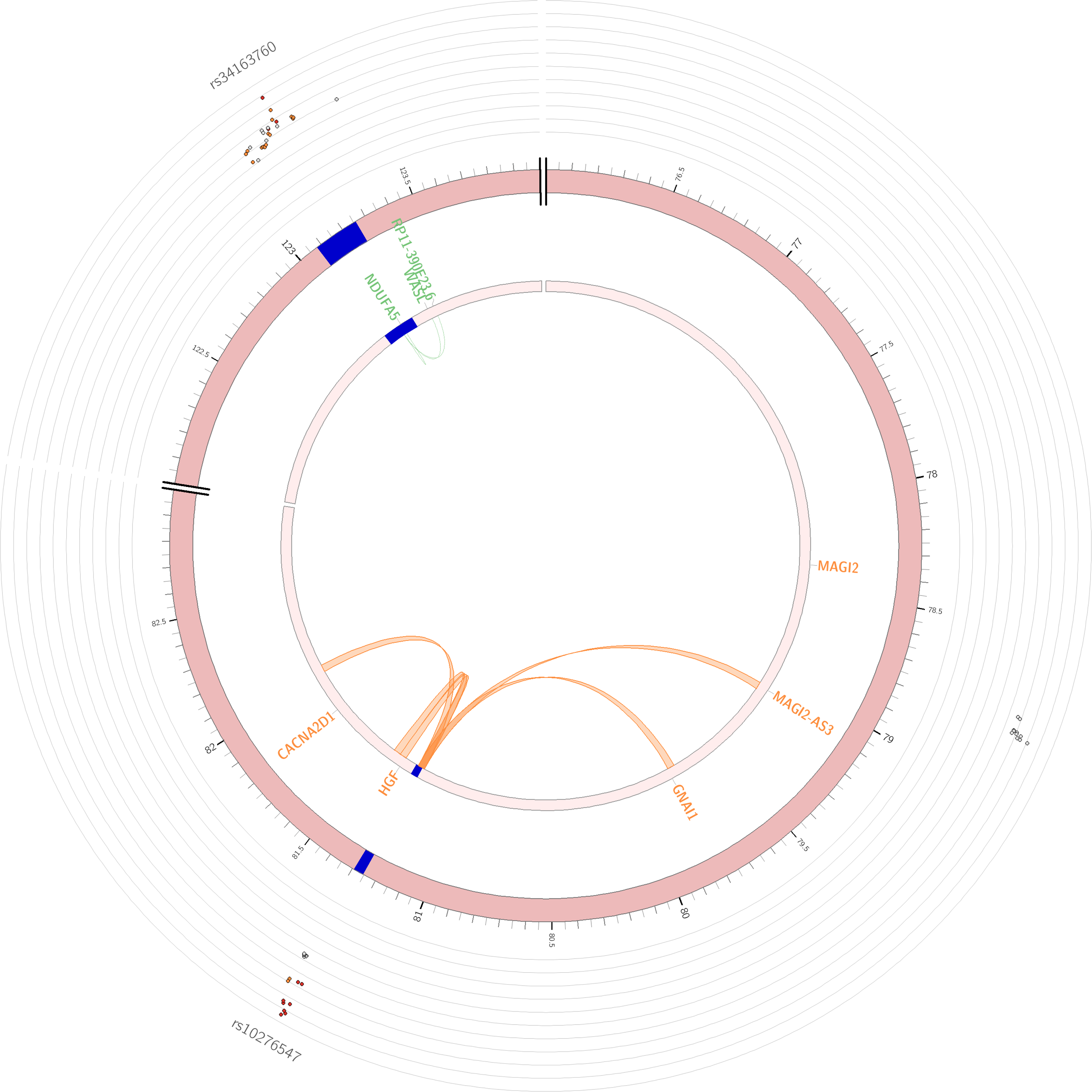
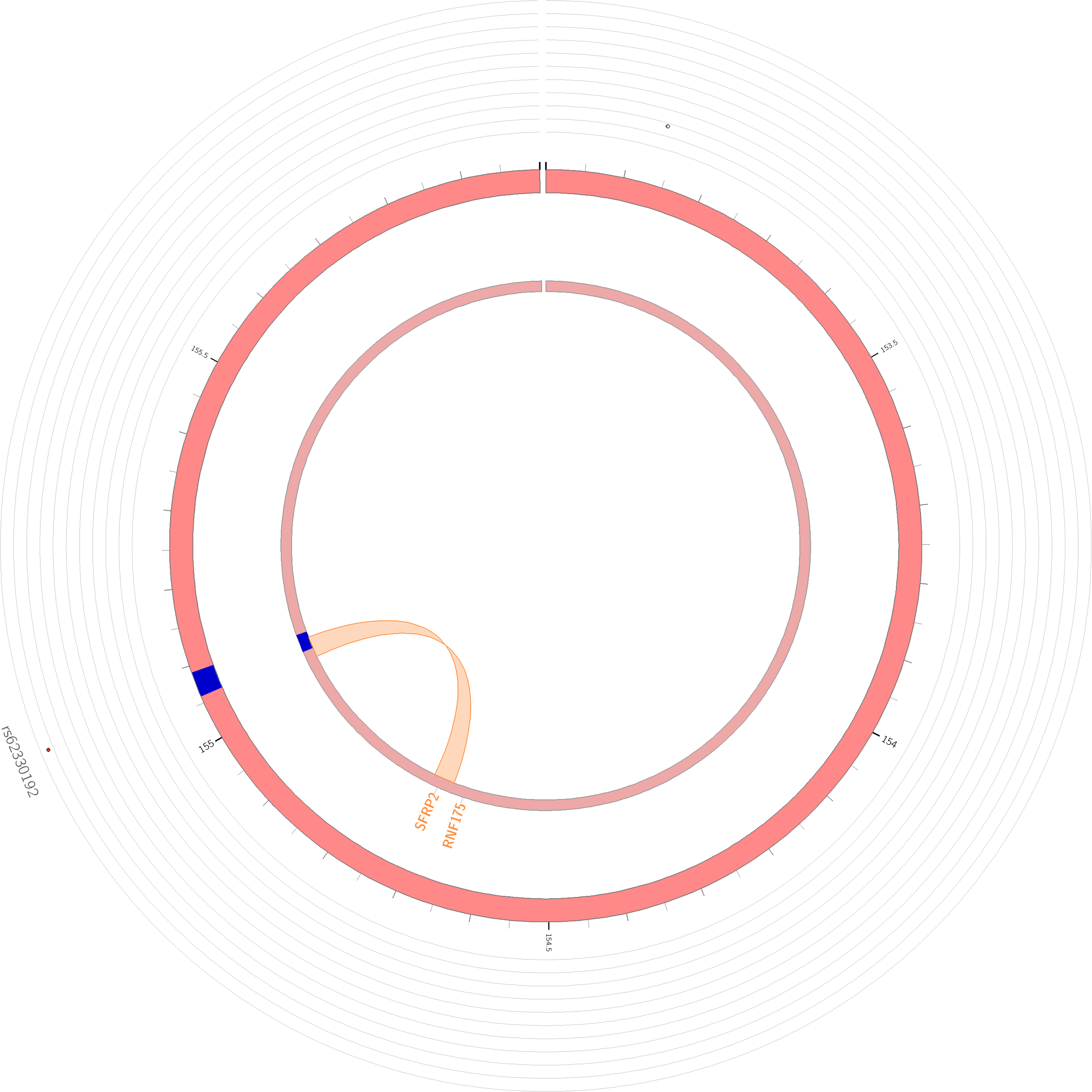
Heat map of 48 prioritized genes based on FUMA related to 12 genomic risk loci in 54 different tissues. The tissue-specific expression patterns are based on GTEx v6 expression data. Cells in red represent a higher expression, compared to the cells in blue. Gene expression comparisons between tissues (horizontal comparison) within a gene (y axis) are comparable but not those of different genes within a tissue (vertical comparison).

**Supplementary Figure S2.**
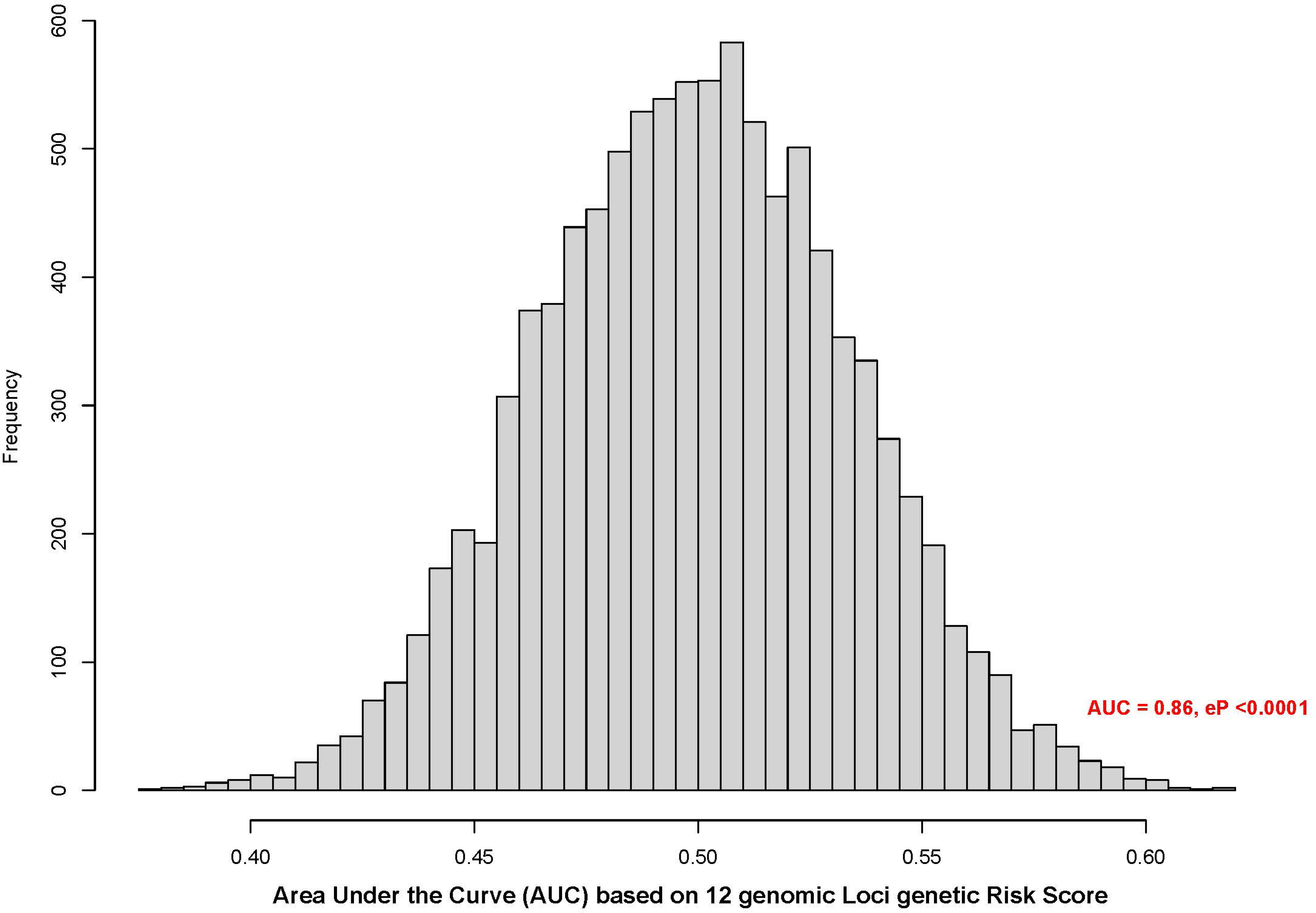
QQ plot of whole genome sequence association analysis of large aneurysm in IVIG-treated Kawasaki Disease. Q-Q plot shows observed distributions of association statistics (-log(p)) in y-axis against those expected (-log(p)) in x-axis under the fitted model.

## Supplementary Tables

**Supplementary Table S1. Independent SNPs (r2 < 0.6) associated with large (medium/giant) aneurysm**

**Supplementary Table S2. Genomic risk loci of interest from large (medium/giant) aneurysm WGS Association**

**Supplementary Table S3. All genes mapped in SNP-based (FUMA) for large (medium/giant) aneurysm**

**Supplementary Table S4. Expression values of the genes prioritized by FUMA for large (medium/giant) aneurysm in 54 tissues**

**Supplementary Table S5. Minor allele frequencies of cases and control design for 4 CAA phenotype**

## REFERENCES

1. McCrindle BW, Rowley AH, Newburger JW, Burns JC, Bolger AF, Gewitz M, Baker AL, Jackson MA, Takahashi M, Shah PB, et al. Diagnosis, Treatment, and Long-Term Management of Kawasaki Disease: A Scientific Statement for Health Professionals From the American Heart Association. Circulation. 2017;135:e927–e999. doi: 10.1161/CIR.0000000000000484

2. Kawasaki T. [Acute febrile mucocutaneous syndrome with lymphoid involvement with specific desquamation of the fingers and toes in children]. Arerugi. 1967;16:178–222.

3. Orenstein JM, Shulman ST, Fox LM, Baker SC, Takahashi M, Bhatti TR, Russo PA, Mierau GW, de Chadarevian JP, Perlman EJ, et al. Three linked vasculopathic processes characterize Kawasaki disease: a light and transmission electron microscopic study. PLoS One. 2012;7:e38998. doi: 10.1371/journal.pone.0038998

4. Kato H, Sugimura T, Akagi T, Sato N, Hashino K, Maeno Y, Kazue T, Eto G, Yamakawa R. Long-term consequences of Kawasaki disease. A 10- to 21-year follow-up study of 594 patients. Circulation. 1996;94:1379-1385.

5. Burns JC, Glode MP. Kawasaki syndrome. Lancet. 2004;364:533–544.

6. Newburger JW, Takahashi M, Beiser AS, Burns JC, Bastian J, Chung KJ, Colan SD, Duffy CE, Fulton DR, Glode MP, et al. A single intravenous infusion of gamma globulin as compared with four infusions in the treatment of acute Kawasaki syndrome. N Engl J Med. 1991;324:1633–1639. doi: 10.1056/NEJM199106063242305

7. Newburger JW, Takahashi M, Burns JC, Beiser AS, Chung KJ, Duffy CE, Glode MP, Mason WH, Reddy V, Sanders SP, et al. The treatment of Kawasaki syndrome with intravenous gamma globulin. N Engl J Med. 1986;315:341–347.

8. Sleeper LA, Minich LL, McCrindle BM, Li JS, Mason W, Colan SD, Atz AM, Printz BF, Baker A, Vetter VL, et al. Evaluation of Kawasaki disease risk-scoring systems for intravenous immunoglobulin resistance. The Journal of pediatrics. 2011;158:831–835 e833. doi: 10.1016/j.jpeds.2010.10.031S0022-3476(10)00922-4 [pii]

9. Newburger JW, Sleeper LA, McCrindle BW, Minich LL, Gersony W, Vetter VL, Atz AM, Li JS, Takahashi M, Baker AL, et al. Randomized trial of pulsed corticosteroid therapy for primary treatment of Kawasaki disease. N Engl J Med. 2007;356:663–675.

10. Miura M, Kobayashi T, Kaneko T, Ayusawa M, Fukazawa R, Fukushima N, Fuse S, Hamaoka K, Hirono K, Kato T, et al. Association of Severity of Coronary Artery Aneurysms in Patients With Kawasaki Disease and Risk of Later Coronary Events. JAMA Pediatr. 2018;172:e180030. doi: 10.1001/jamapediatrics.2018.0030

11. McCrindle BW, Manlhiot C, Newburger JW, Harahsheh AS, Giglia TM, Dallaire F, Friedman K, Low T, Runeckles K, Mathew M, et al. Medium-Term Complications Associated With Coronary Artery Aneurysms After Kawasaki Disease: A Study From the International Kawasaki Disease Registry. J Am Heart Assoc. 2020;9:e016440. doi: 10.1161/JAHA.119.016440

12. Friedman KG, Gauvreau K, Hamaoka-Okamoto A, Tang A, Berry E, Tremoulet AH, Mahavadi VS, Baker A, deFerranti SD, Fulton DR, et al. Coronary Artery Aneurysms in Kawasaki Disease: Risk Factors for Progressive Disease and Adverse Cardiac Events in the US Population. J Am Heart Assoc. 2016;5. doi: 10.1161/JAHA.116.003289

13. Patel AS, Bruce M, Harrington W, Portman MA. Coronary artery stenosis risk and time course in Kawasaki disease patients: experience at a US tertiary pediatric centre. Open Heart. 2015;2:e000206. doi: 10.1136/openhrt-2014-000206

14. Newburger JW, Taubert KA, Shulman ST, Rowley AH, Gewitz MH, Takahashi M, McCrindle BW. Summary and abstracts of the Seventh International Kawasaki Disease Symposium: December 4–7, 2001, Hakone, Japan. Pediatr Res. 2003;53:153-157.

15. Lee JJY, Lin E, Widdifield J, Mahood Q, McCrindle BW, Yeung RSM, Feldman BM. The Long-term Cardiac and Noncardiac Prognosis of Kawasaki Disease: A Systematic Review. Pediatrics. 2022;149. doi: 10.1542/peds.2021-052567

16. Hu J, Ren W. Analysis of the Risk Factors in Prognosis of Kawasaki Disease With Coronary Artery Lesions. Front Pediatr. 2021;9:798148. doi: 10.3389/fped.2021.798148

17. Lin MT, Sun LC, Wu ET, Wang JK, Lue HC, Wu MH. Acute and late coronary outcomes in 1073 patients with Kawasaki disease with and without intravenous gamma-immunoglobulin therapy. Arch Dis Child. 2015;100:542–547. doi: 10.1136/archdischild-2014-306427

18. Kobayashi T, Inoue Y, Otani T, Morikawa A, Kobayashi T, Takeuchi K, Saji T, Sonobe T, Ogawa S, Miura M, et al. Risk stratification in the decision to include prednisolone with intravenous immunoglobulin in primary therapy of Kawasaki disease. Pediatr Infect Dis J. 2009;28:498–502.

19. Davies S, Sutton N, Blackstock S, Gormley S, Hoggart CJ, Levin M, Herberg JA. Predicting IVIG resistance in UK Kawasaki disease. Arch Dis Child. 2015;100:366–368. doi: 10.1136/archdischild-2014-307397

20. Fu PP, Du ZD, Pan YS. Novel predictors of intravenous immunoglobulin resistance in Chinese children with Kawasaki disease. Pediatr Infect Dis J. 2013;32:e319–323. doi: 10.1097/INF.0b013e31828e887f

21. Shin J, Lee H, Eun L. Verification of Current Risk Scores for Kawasaki Disease in Korean Children. J Korean Med Sci. 2017;32:1991–1996. doi: 10.3346/jkms.2017.32.12.1991

22. Portman MA, Shrestha S. One Size Does Not Fit All: Genetic Prediction of Kawasaki Disease Treatment Response in Diverse Populations. Circulation Cardiovascular genetics. 2017;10. doi: 10.1161/CIRCGENETICS.117.001917

23. Makowsky R, Wiener HW, Ptacek TS, Silva M, Shendre A, Edberg JC, Portman MA, Shrestha S. FcgammaR gene copy number in Kawasaki disease and intravenous immunoglobulin treatment response. Pharmacogenet Genomics. 2013;23:455–462. doi: 10.1097/FPC.0b013e328363686e

24. Shrestha S, Wiener H, Shendre A, Kaslow RA, Wu J, Olson A, Bowles NE, Patel H, Edberg JC, Portman MA. Role of activating FcgammaR gene polymorphisms in Kawasaki disease susceptibility and intravenous immunoglobulin response. Circulation Cardiovascular genetics. 2012;5:309–316. doi: 10.1161/CIRCGENETICS.111.962464

25. Shrestha S, Wiener HW, Olson AK, Edberg JC, Bowles NE, Patel H, Portman MA. Functional FCGR2B gene variants influence intravenous immunoglobulin response in patients with Kawasaki disease. The Journal of allergy and clinical immunology. 2011;128:677–680. doi: 10.1016/j.jaci.2011.04.027

26. Wu SF, Chang JS, Wan L, Tsai CH, Tsai FJ. Association of IL-1Ra gene polymorphism, but no association of IL-1beta and IL-4 gene polymorphisms, with Kawasaki disease. J Clin Lab Anal. 2005;19:99–102.

27. Minami T, Suzuki H, Takeuchi T, Uemura S, Sugatani J, Yoshikawa N. A polymorphism in plasma platelet-activating factor acetylhydrolase is involved in resistance to immunoglobulin treatment in Kawasaki disease. The Journal of pediatrics. 2005;147:78–83.

28. Burns JC, Shimizu C, Shike H, Newburger JW, Sundel RP, Baker AL, Matsubara T, Ishikawa Y, Brophy VA, Cheng S, et al. Family-based association analysis implicates IL-4 in susceptibility to Kawasaki disease. Genes Immun. 2005;6:438–444.

29. Burns JC, Shimizu C, Gonzalez E, Kulkarni H, Patel S, Shike H, Sundel RS, Newburger JW, Ahuja SK. Genetic Variations in the Receptor-Ligand Pair CCR5 and CCL3L1 Are Important Determinants of Susceptibility to Kawasaki Disease. J Infect Dis. 2005;192:344–349.

30. Khor CC, Davila S, Breunis WB, Lee YC, Shimizu C, Wright VJ, Yeung RS, Tan DE, Sim KS, Wang JJ, et al. Genome-wide association study identifies FCGR2A as a susceptibility locus for Kawasaki disease. Nat Genet. 2011;43:1241–1246. doi: 10.1038/ng.981

31. Onouchi Y, Ozaki K, Burns JC, Shimizu C, Terai M, Hamada H, Honda T, Suzuki H, Suenaga T, Takeuchi T, et al. A genome-wide association study identifies three new risk loci for Kawasaki disease. Nat Genet. 2012;44:517–521. doi: 10.1038/ng.2220

32. Holman RC, Christensen KY, Belay ED, Steiner CA, Effler PV, Miyamura J, Forbes S, Schonberger LB, Melish M. Racial/ethnic differences in the incidence of Kawasaki syndrome among children in Hawaii. Hawaii medical journal. 2010;69:194–197.

33. Padilla LA, Collins JL, Idigo AJ, Lau Y, Portman MA, Shrestha S. Kawasaki Disease and Clinical Outcome Disparities Among Black Children. The Journal of pediatrics. 2021;229:54–60 e52. doi: 10.1016/j.jpeds.2020.09.052

34. Shrestha S, Wiener HW, Kajimoto H, Srinivasasainagendra V, Ledee D, Chowdhury S, Cui J, Chen JY, Beckley MA, Padilla LA, et al. Pharmacogenomics of intravenous immunoglobulin response in Kawasaki disease. Frontiers in Immunology. 2024;14. doi: 10.3389/fimmu.2023.1287094

35. Newburger JW, Takahashi M, Gerber MA, Gewitz MH, Tani LY, Burns JC, Shulman ST, Bolger AF, Ferrieri P, Baltimore RS, et al. Diagnosis, treatment, and long-term management of Kawasaki disease: a statement for health professionals from the Committee on Rheumatic Fever, Endocarditis, and Kawasaki Disease, Council on Cardiovascular Disease in the Young, American Heart Association. Pediatrics. 2004;114:1708–1733.

36. Watanabe K, Taskesen E, van Bochoven A, Posthuma D. Functional mapping and annotation of genetic associations with FUMA. Nat Commun. 2017;8:1826. doi: 10.1038/s41467-017-01261-5

37. Consortium GT. Human genomics. The Genotype-Tissue Expression (GTEx) pilot analysis: multitissue gene regulation in humans. Science. 2015;348:648–660. doi: 10.1126/science.1262110

38. van Berkum NL, Lieberman-Aiden E, Williams L, Imakaev M, Gnirke A, Mirny LA, Dekker J, Lander ES. Hi-C: a method to study the three-dimensional architecture of genomes. J Vis Exp. 2010. doi: 10.3791/1869

39. Kircher M, Witten DM, Jain P, O’Roak BJ, Cooper GM, Shendure J. A general framework for estimating the relative pathogenicity of human genetic variants. Nat Genet. 2014;46:310–315. doi: 10.1038/ng.2892

40. Roadmap Epigenomics C, Kundaje A, Meuleman W, Ernst J, Bilenky M, Yen A, Heravi-Moussavi A, Kheradpour P, Zhang Z, Wang J, et al. Integrative analysis of 111 reference human epigenomes. Nature. 2015;518:317–330. doi: 10.1038/nature14248

41. Herberman RB, Nunn ME, Holden HT, Lavrin DH. Natural cytotoxic reactivity of mouse lymphoid cells against syngeneic and allogeneic tumors. II. Characterization of effector cells. Int J Cancer. 1975;16:230–239. doi: 10.1002/ijc.2910160205

42. Kiessling R, Klein E, Wigzell H. "Natural" killer cells in the mouse. I. Cytotoxic cells with specificity for mouse Moloney leukemia cells. Specificity and distribution according to genotype. Eur J Immunol. 1975;5:112–117. doi: 10.1002/eji.1830050208

43. Trinchieri G, Santoli D. Anti-viral activity induced by culturing lymphocytes with tumor-derived or virus-transformed cells. Enhancement of human natural killer cell activity by interferon and antagonistic inhibition of susceptibility of target cells to lysis. J Exp Med. 1978;147:1314–1333. doi: 10.1084/jem.147.5.1314

44. Long EO, Kim HS, Liu D, Peterson ME, Rajagopalan S. Controlling natural killer cell responses: integration of signals for activation and inhibition. Annu Rev Immunol. 2013;31:227–258. doi: 10.1146/annurev-immunol-020711-075005

45. Choi IS, Lee MJ, Choi SA, Choi KS, Jeong IS, Cho HJ. Circulating Immune Cell Profile and Changes in Intravenous Immunoglobulin Responsiveness Over the Disease Course in Children With Kawasaki Disease. Front Pediatr. 2021;9:792870. doi: 10.3389/fped.2021.792870

46. Cheong KX, Yong RYY, Tan MMH, Tey FLK, Ang BCH. Association of VIPR2 and ZMAT4 with high myopia. Ophthalmic Genet. 2020;41:41–48. doi: 10.1080/13816810.2020.1737951

47. Wan J, Gao Y, Zhao X, Wu Q, Fu X, Shao Y, Yang H, Guan M, Yu B, Zhang W. The association between the copy-number variations of ZMAT4 and hematological malignancy. Hematology. 2011;16:20–23. doi: 10.1179/102453311X12902908411751

48. Helms AS, Tang VT, O’Leary TS, Friedline S, Wauchope M, Arora A, Wasserman AH, Smith ED, Lee LM, Wen XW, et al. Effects of MYBPC3 loss-of-function mutations preceding hypertrophic cardiomyopathy. JCI Insight. 2020;5. doi: 10.1172/jci.insight.133782

49. van Velzen HG, Schinkel AFL, Oldenburg RA, van Slegtenhorst MA, Frohn-Mulder IME, van der Velden J, Michels M. Clinical Characteristics and Long-Term Outcome of Hypertrophic Cardiomyopathy in Individuals With a MYBPC3 (Myosin-Binding Protein C) Founder Mutation. Circulation Cardiovascular genetics. 2017;10. doi: 10.1161/CIRCGENETICS.116.001660

50. Al-Khelaifi F, Yousri NA, Diboun I, Semenova EA, Kostryukova ES, Kulemin NA, Borisov OV, Andryushchenko LB, Larin AK, Generozov EV, et al. Genome-Wide Association Study Reveals a Novel Association Between MYBPC3 Gene Polymorphism, Endurance Athlete Status, Aerobic Capacity and Steroid Metabolism. Front Genet. 2020;11:595. doi: 10.3389/fgene.2020.00595

51. Kuo HC, Li SC, Guo MM, Huang YH, Yu HR, Huang FC, Jiao F, Kuo HC, Andrade J, Chan WC. Genome-Wide Association Study Identifies Novel Susceptibility Genes Associated with Coronary Artery Aneurysm Formation in Kawasaki Disease. PLoS One. 2016;11:e0154943. doi: 10.1371/journal.pone.0154943

52. Beckley MA, Shrestha S, Singh KK, Portman MA. The role of mitochondria in the pathogenesis of Kawasaki disease. Front Immunol. 2022;13:1017401. doi: 10.3389/fimmu.2022.1017401

53. Hoggart C, Shimizu C, Galassini R, Wright VJ, Shailes H, Bellos E, Herberg JA, Pollard AJ, O’Connor D, Choi SW, et al. Identification of novel locus associated with coronary artery aneurysms and validation of loci for susceptibility to Kawasaki disease. Eur J Hum Genet. 2021;29:1734–1744. doi: 10.1038/s41431-021-00838-5

54. Kim JJ, Park YM, Yoon D, Lee KY, Seob Song M, Doo Lee H, Kim KJ, Park IS, Nam HK, Weon Yun S, et al. Identification of KCNN2 as a susceptibility locus for coronary artery aneurysms in Kawasaki disease using genome-wide association analysis. J Hum Genet. 2013;58:521–525. doi: 10.1038/jhg.2013.43

55. Liu J, Yuan P, Pang Y, Su D. ITPKC polymorphism (rs7251246 T > C), coronary artery aneurysms, and thrombosis in patients with Kawasaki disease in a Southern Han Chinese population. Front Immunol. 2023;14:1184162. doi: 10.3389/fimmu.2023.1184162

56. Oates-Whitehead RM, Baumer JH, Haines L, Love S, Maconochie IK, Gupta A, Roman K, Dua JS, Flynn I. Intravenous immunoglobulin for the treatment of Kawasaki disease in children. Cochrane Database Syst Rev. 2003;2003:CD004000. doi: 10.1002/14651858.CD004000

57. Shimizu C, Eleftherohorinou H, Wright VJ, Kim J, Alphonse MP, Perry JC, Cimaz R, Burgner D, Dahdah N, Hoang LT, et al. Genetic Variation in the SLC8A1 Calcium Signaling Pathway Is Associated With Susceptibility to Kawasaki Disease and Coronary Artery Abnormalities. Circulation Cardiovascular genetics. 2016;9:559–568. doi: 10.1161/CIRCGENETICS.116.001533

58. Shimizu C, Matsubara T, Onouchi Y, Jain S, Sun S, Nievergelt CM, Shike H, Brophy VH, Takegawa T, Furukawa S, et al. Matrix metalloproteinase haplotypes associated with coronary artery aneurysm formation in patients with Kawasaki disease. J Hum Genet. 2010;55:779–784. doi: 10.1038/jhg.2010.109

59. Kwon YC, Kim JJ, Yu JJ, Yun SW, Yoon KL, Lee KY, Kil HR, Kim GB, Han MK, Song MS, et al. Identification of the TIFAB Gene as a Susceptibility Locus for Coronary Artery Aneurysm in Patients with Kawasaki Disease. Pediatr Cardiol. 2019;40:483–488. doi: 10.1007/s00246-018-1992-7

60. Lin YJ, Chang JS, Liu X, Tsang H, Chien WK, Chen JH, Hsieh HY, Hsueh KC, Shiao YT, Li JP, et al. Genetic variants in PLCB4/PLCB1 as susceptibility loci for coronary artery aneurysm formation in Kawasaki disease in Han Chinese in Taiwan. Sci Rep. 2015;5:14762. doi: 10.1038/srep14762

61. Wu J, Pfeiffer RM, Gail MH. Strategies for developing prediction models from genome-wide association studies. Genet Epidemiol. 2013;37:768–777. doi: 10.1002/gepi.21762

